# Overall vertical transmission of HCV, transmission net of clearance, and timing of transmission

**DOI:** 10.1101/2021.09.28.21264075

**Authors:** A E Ades, Fabiana Gordon, Karen Scott, Jeannie Collins, Claire Thorne, Lucy Pembrey, Elizabeth Chappell, Eugènia Mariné-Barjoan, Karina Butler, Giuseppe Indolfi, Diana M Gibb, Ali Judd

## Abstract

**Background:** It is widely accepted that the risk of HCV vertical transmission (VT) is 5-6% in mono-infected women, and that 25-40% of HCV infection clears spontaneously within 5 years. However, VT and clearance rates have not been estimated from the same datasets, and there is a lack of information on VT rates “net” of clearance.

**Methods:** We re-analysed data on 1749 children in 3 prospective cohorts to obtain coherent estimates of overall VT rate and VT rates “net” of clearance at different ages. Clearance rates were used to impute the proportion of uninfected children who had been infected and then cleared before testing negative. The proportion of transmission early in utero, late in utero and at delivery was estimated from data on the proportion of RNA positives in samples tested within three days of birth, and differences between elective caesarean and non-elective caesarean deliveries.

**Findings:** Overall VT rates were 7.2% (95% credible interval 5.6-8.9) in mothers who were HIV negative and 12.1% (8.6-16.8) in HIV-co-infected women. The corresponding rates net of clearance at 5 years were 2.4% (1.1-4.1) and 4.1% (1.7-7.3). We estimated that 24.8% (12.1-40.8) of infections occur early in utero, 66.0% (42.5-83.3) later in utero, and 9.3% (0.5-30.6) during delivery.

**Conclusion:** Overall VT rates are about 24% higher than previously assumed, but the risk of infection persisting beyond age 5 years is about 38% lower. The results can inform design of trials of to prevent or treat pediatric HCV infection, and strategies to manage children exposed in utero.

**Key points:** Taking account of infections that would have cleared spontaneously before detection, the rate of HCV vertical transmission is 7.2% (95%CrI 5.6-8.9) in mono-infected women, but transmission “net” of clearance is 3.1% (1.8-4.4) at 3 years, and 2.4% (1.1-4.1) at 5.

---

With the discovery of direct acting antivirals (DAAs) to treat hepatitis C virus (HCV), attention is turning to interventions either in pregnancy or in infancy to prevent or treat vertically acquired infection. The WHO’s target of HCV elimination by 2030[1] has added further urgency to this issue. According to a 2014 meta-analysis vertical transmission (VT) occurs in 5.8% of infants of HCV-RNA positive mothers who are not HIV-co-infected, and 10.8% if mothers also have HIV.[2] High HCV viral load, and prolonged rupture of membranes have also been associated with higher risk.[3]

The VT and clearance rates reported in the literature depend not only on how pediatric infection is defined, but also on the timing of diagnostic tests. Previous investigators have commented on the lack of standardization in testing schedules and in methods for calculating transmission rates.[21, 22] To illustrate, consider an infant whose first RNA test is at 3 months and is negative. This infant would be counted as uninfected in a prospective study estimating the VT rate. However, there is a possibility that the infant had been infected but that the infection cleared before 3 months. Moreover, if the first negative RNA test was at 6 months, an initial infection would have had longer in which to clear, and the possibility that the child had originally been infected would be correspondingly greater.

Infections that clear within the first 3 or 6 months do not require treatment or prevention, as it is assumed that they have no clinical significance. Even so, if it is true that unobserved spontaneous clearance is occurring, then VT rates calculated by the usual methods must be under-estimates. This introduces an undesirable lack of clarity about how much clearance might already be “factored in” to the commonly cited 5-10% transmission rates.

Unfortunately, very few studies have estimated both VT and clearance rates in the exact same dataset.[22, 23] A single, coherent, account of the underlying VT rate and the VT rate net of clearance at different ages is needed to inform strategies for prevention, diagnosis, and treatment of vertically-acquired infection, and are especially important for planning trials of preventive and therapeutic interventions. Information on the timing and mechanisms of transmission may also be helpful in determining the optimal timing of preventive treatment in pregnancy.

In this paper, we use data on individual mother-child pairs from three published European cohorts to estimate, for the first time, both the overall rate of confirmed VT and the VT rates “net” of clearance at ages between 6 months and 5 years. The clearance rates assumed in this study are those relating to confirmed infection, estimated previously from the same data.[24] The analysis described here looks at a limited number of risk factors, mother’s HCV-RNA viral load, mother’s HIV coinfection, and mode of delivery. We also investigate the timing and mechanism of infection, by estimating the proportion of infection that occurs early in utero, later in utero and during delivery.

## METHODS

### Data sources

Three prospective studies following infants born to HCV anti-body positive mothers were included: European Pediatric HCV Network (EPHN);[3, 8, 25, 26] the British Paediatric Surveillance Unit (BPSU) study, which included 3 hospitals in Dublin, Irish Republic and centres across the UK;[10] and the ALHICE study (Alpes-Maritimes, Languedoc, Haute Garonne Infection C chez l’Enfant).[27] The process by which these studies were selected has been described previously.[24]

### Definitions

Infants were regarded as *Infected* if they were anti-HCV positive after 18 months and/or had at least two positive RNA tests; otherwise they were *Uninfected* if they tested RNA negative at least once after age 6 weeks or if their final anti-HCV test was negative; otherwise they were *Indeterminate*. Note that “Infected” is to be interpreted as “ever-infected” because it is recognized that infected infants can clear infection, and that “uninfected” infants may have been infected and cleared. Supporting details are given in Supplementary Materials.

Ages at which tests are performed play a key role in the analysis. In Indeterminate infants we define *Age at last anti-HCV positive under 18 months:* the later the last positive anti-HCV test, the more likely the infant is to be infected. We also define *Age at last RNA negative under 6 weeks:* the later this is the less likely the infant is to have been infected. In Uninfected infants we also define *Age at last RNA negative under 6 weeks*, and in addition *Age at first anti-HCV negative test or the first negative RNA test over 6 weeks, whichever is earliest*. This is age when the infant is first known to be uninfected: the later this is the more likely the infant is to have been infected and cleared.

### Statistical methods

Our objective was to estimate the risk of vertical infection, the impact of risk factors (mother’s HIV and HCV-RNA viral load), and the proportions of infection transmitted Early in Utero (EiU), Late in Utero (LiU) and at delivery. The proportion transmitted EiU is informed directly by the proportion RNA positive in the first 3 days. Assuming that children delivered by elective caesarean cannot acquire infection during delivery, the difference between overall transmission rates in ECS and non-ECS modes of delivery informs the proportion of non-EiU transmission that is LiU as opposed to occurring during delivery, among those not delivered by ECS.

The underlying model assumes that infants can be infected EiU, LiU or during delivery, with infection at each stage being conditional on not being infected at an earlier stage. Data is available on risk factors (Study: EPHN, BPSU, ALHICE; mother’s HIV status; and mother’s HCV viral load measured as near as possible to delivery: Low, High (>600 copies/ml)). Risk factors impact on risk of transmission in each of the three routes as they would in a standard logistic regression, but it is assumed the impact is the same for each route. We assumed that there could be an interaction between mother’s HIV and HCV-RNA viral load, so that the log odds ratio associated viral load could depend on HIV status. However, we set this constrained interaction model so that the log risk attached to HIV and high HCV viral load combined had to be greater than the risk of either factor alone, but no more than both added together. Standard interaction and main effect models were investigated as sensitivity analyses. All models controlled for study effects.

To simplify the analysis, mother-child pairs lacking data on mode of delivery or mother’s HIV-status, and cases where the mother was known to be HCV-RNA negative were excluded. Mother’s RNA status was unknown in 67% of the remaining records, and where RNA status was known to be positive, HCV viral load was unknown in 43% (Table 1). To make best use of data available we assumed that the proportions of mothers with low viral load, or no detectable RNA, stratified by both mother’s HIV status and mode of delivery were exactly the same as where this information was known in the EPHN study. We assessed robustness of conclusions to these assumptions by sensitivity analyses assuming higher or lower proportions with low viral load and with no HCV RNA.

**Table 1.** Infection status and risk factor distribution in the three cohorts, after removal of records with missing HIV, missing mode of delivery, and RNA-negative mothers. NK: not known. ECS: Elective caesarean section. RNA+: HCV RNA positive. Mean and 95% CrI.

|  |  | EPHN |  | BPSU |  | ALHICE |  | TOTAL |  |
| --- | --- | --- | --- | --- | --- | --- | --- | --- | --- |
|  |  | n | % | n | % | n | % | n | % |
| TOTAL |  | 1256 | 100 | 342 | 100 | 151 | 100 | 1749 | 100 |
| Infection status | Infected | 69 | 5.5 | 15 | 4.4 | 12 | 7.9 | 96 | 5.5 |
|  | Indeterminate | 121 | 9.6 | 102 | 29.8 | 0 | 0.0 | 223 | 12.8 |
|  | Uninfected | 1066 | 84.9 | 225 | 65.8 | 139 | 92.1 | 1430 | 81.8 |
| Mother's HIV | No | 1053 | 83.8 | 321 | 93.1 | 105 | 69.5 | 1479 | 84.6 |
|  | Yes | 203 | 16.2 | 21 | 6.9 | 46 | 30.5 | 270 | 15.4 |
| Mother's HCV Viral Load | Low | 167 | 13.3 | 0 | 0.0 | 94 | 62.3 | 261 | 14.9 |
|  | High | 29 | 2.3 | 0 | 0.0 | 39 | 25.8 | 68 | 3.9 |
|  | NK but RNA+ | 240 | 19.1 | 0 | 0.0 | 4 | 2.6 | 244 | 14.0 |
|  | RNA NK | 820 | 65.3 | 342 | 100 | 14 | 9.3 | 1176 | 67.2 |
| Mode of Delivery | ECS | 373 | 29.7 | 26 | 3.3 | 35 | 23.2 | 434 | 24.8 |
|  | Non-ECS | 883 | 70.3 | 316 | 96.7 | 116 | 76.8 | 1315 | 75.2 |

In outline, the statistical analysis estimates the probability that each child of indeterminate status is infected, based on their risk group and the age when they were last anti-HCV positive and the age at the last HCV-RNA negative if this was under 6 weeks. Similarly, the probability that each uninfected child was originally infected and then cleared is calculated, based again on risk group, and on the age when they were first ascertained as uninfected. The basis for these probability calculations is shown in Supplementary materials Table S1. The probabilities of infection in indeterminate and uninfected children are then added together, and added to the number of children with confirmed infection to form a notional transmission rate

To estimate the overall VT rate, the analytic method in a sense “restores” the proportion of uninfected children who were originally infected but then cleared, based on the rate of clearance and age when they were first known to be negative, and then, to estimate the net VT rates at selected ages, it “removes” them assuming the same clearance rate.

The statistical analysis was carried out using Bayesian Markov Chain Monte Carlo estimation. Details of the statistical methods are given in the Supplementary Materials.

## RESULTS

The proportions infected, indeterminate and uninfected and the risk factor distributions are shown in Table 1.

Figure 1 panel A1 shows the probability that uninfected children were anti-HCV positive by age; B1 the probability that infected children were RNA negative under 6 weeks of age; C1 the probability that an infected child had not cleared by age. These functions had been estimated from the three cohorts in advance. Panel A2 is a histogram showing age at last positive anti-HCV among children with indeterminate status; B2 shows age at last RNA negative under 6 weeks in uninfected and indeterminate children; C2 shows age at first RNA or anti-HCV negative among uninfected children. The mean age when uninfected children of mono-infected women were first known to be uninfected was 5.2 months in, and 4.4 months in HIV co-infected women

**Figure 1.**
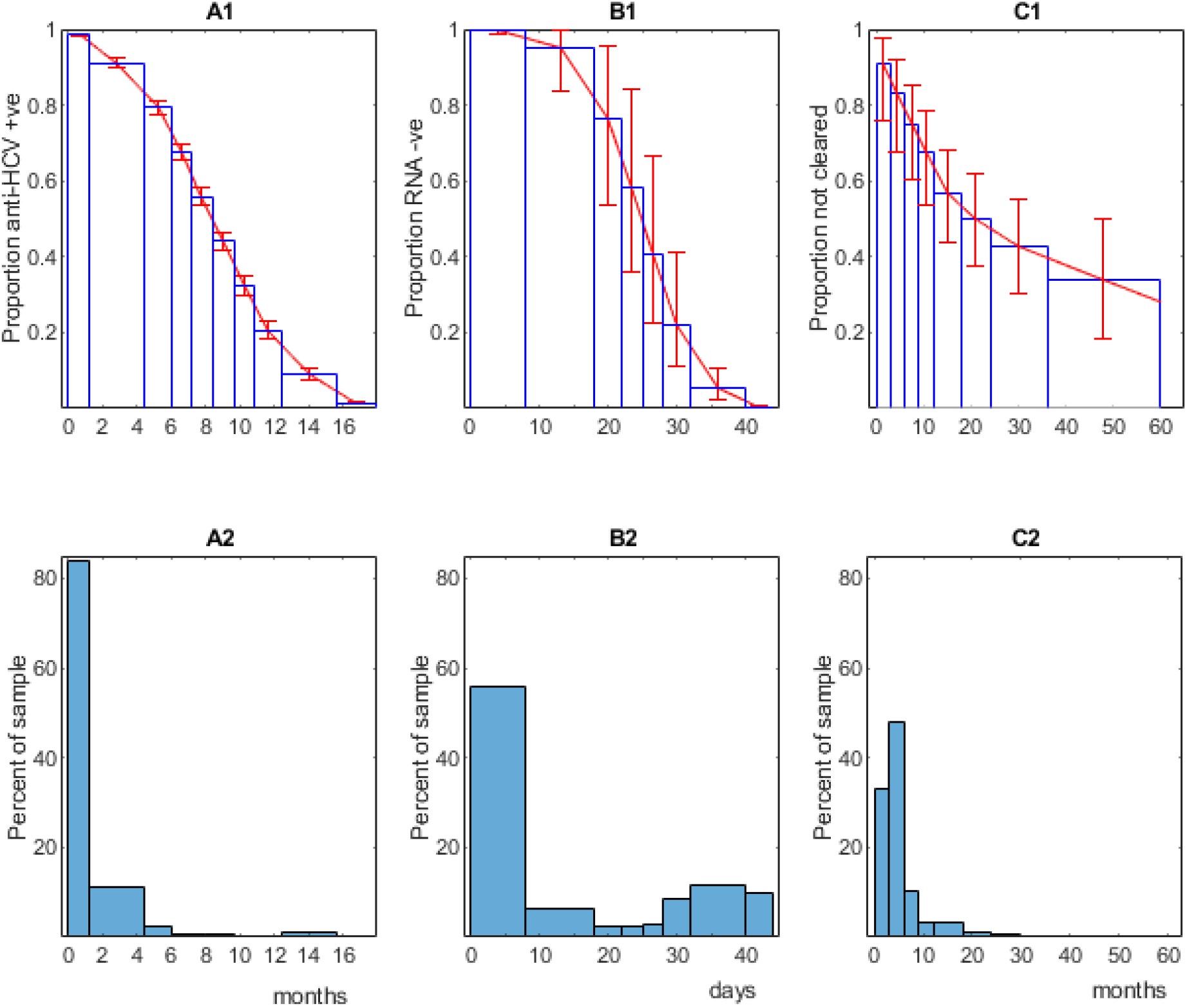
Panel **A1**. Assumed proportion of uninfected children remaining anti-HCV positive at each age up to 18 months. **B1** Assumed proportion of infected children who are not initially RNA positive remaining RNA negative by age up to 6 weeks. **C1** proportion of infection not yet cleared, by age up to 5 years. **A2** proportion of indeterminates with last anti-HIV +ve at each age. **B2** proportion of uninfected and indeterminate with last RNA negative at each age (< 6 weeks). **C2** proportion of uninfected children with the first test indicating they were uninfected at each age.

Table 2 illustrates the results of imputing the probability of infection in each indeterminate and uninfected child. In addition to the 96 observed infections, there were a further estimated 10.2 infections among the 223 children with indeterminate status, and a further 9.0 unobserved infections among the 1430 nominally uninfected infants. In all, there were therefore an estimated total 115.2 infections. The VT rate among the uninfected is under 1%, but the imputed 9.0 “missed” infections represents 7.8% of the total 115.2. In the entire combined cohort of 1749, there is a nominal VT rate of 6.6% (6.2 – 7.1) (Table 2). Note, however, that the study population included 67% mothers who were anti-HCV positive but with unknown HCV-RNA status, a proportion of whom would have been RNA negative and would not have transmitted.

**Table 2.** Observed and unobserved infections, nominal overall vertical transmission rates.

|  | Infected | Indeterminate | Uninfected | Total |
| --- | --- | --- | --- | --- |
| Totals | 96 | 223 | 1430 | 1739 |
| Observed infections | 96 | - | - | 115.2 (108.7-124.2) |
| Unobserved infections | - | 10.2 (7.1 – 13.8) | 9.0 (4.3-17.1) |  |
| VT rate % | - | 4.7 (3.2-6.3) | 0.6 (0.3-1.2) | 6.6 (6.2-7.1) |

Analysis of risk factors (Table 3) suggests no important differences between studies, and strong effects of both maternal HIV status and maternal HCV-RNA viral load. Also shown are the absolute risks of transmission at each stage: early in utero, late in utero and at delivery, in the HIV negative low HCV-RNA viral load group. The proportion of transmission by each route (Table 4) indicates that in non-ECS deliveries, 24.8, 66.0 and 9.3 percent of transmissions occur early in utero, late in utero and at delivery. Among ECS deliveries we estimated 27.5% early and 72.5 late in utero. However, credible intervals were wide.

**Table 3.** Risk of vertical transmission by route, and odds ratios for study and risk group, median and 95% CrI.

|  | Median | 2.5% | 97.5% |
| --- | --- | --- | --- |
| **Risk of transmission, %, by route** |  |  |  |
| Early in utero | 1.38 | 0.64 | 2.59 |
| Late in utero | 3.88 | 2.24 | 5.87 |
| Delivery | 0.38 | 0.03 | 1.97 |
| **Odds ratios: study** |  |  |  |
| EPHN | 1 (ref) | - | - |
| BPSU | 1.23 | 0.64 | 2.20 |
| ALHICE | 0.98 | 0.47 | 1.87 |
| **Odds ratios: risk group** |  |  |  |
| HIV-, Low VL | 1 (ref) | - | - |
| HIV-, High VL | 2.66 | 1.19 | 6.12 |
| HIV+, Low VL | 1.75 | 1.08 | 3.12 |
| HIV+, High VL | 3.43 | 1.69 | 8.03 |

**Table 4.**
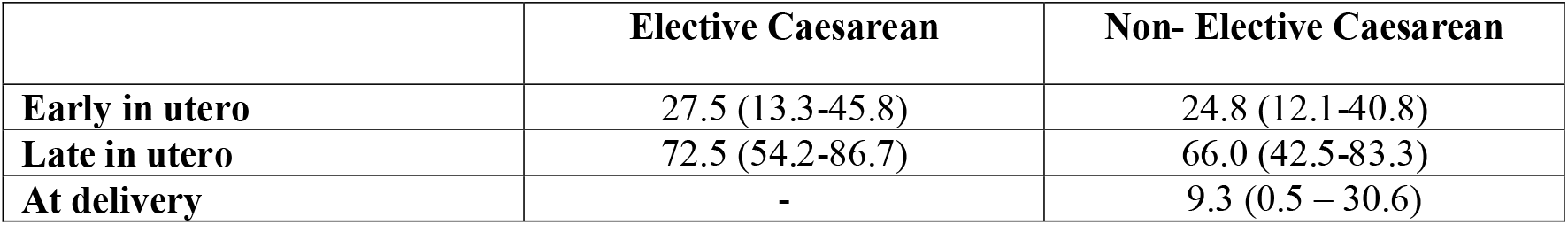
Percent of vertical infection by stage, mean and 95% CrI.

The overall VT rates by maternal HIV status, HCV viral load and mode of delivery are shown in Table 5, and the average *net* VT rates at ages from 3 months to 5 years are plotted in Figure 2 separately for children of mono-infected and HIV-co-infected mothers. In these groups overall transmission risks are 7.2% and 12.1% respectively, falling to VT rates net of clearance at 5 years of 2.4% (1.1-4.1) and 4.1% (1.7-7.3).

**Table 5.** Overall VT rates, by subgroup.

| Mode of delivery | HCV Viral load | HIV -ve |  |  | HIV +ve |  |  |
| --- | --- | --- | --- | --- | --- | --- | --- |
|  |  | mean | 2.5% | 97.5% | mean | 2.5% | 97.5% |
| ECS | Low | 5.6 | 3.7 | 7.6 | 9.6 | 5.6 | 14.8 |
|  | High | 14.2 | 7.1 | 23.5 | 17.5 | 10.2 | 27.7 |
| Non-ECS | Low | 6.1 | 4.2 | 8.2 | 10.6 | 6.2 | 16.2 |
|  | High | 15.3 | 8.1 | 24.6 | 19.1 | 11.4 | 29.8 |
| Weighted average |  | 7.2 | 5.6 | 8.9 | 12.1 | 8.6 | 16.8 |

**Figure 2.**
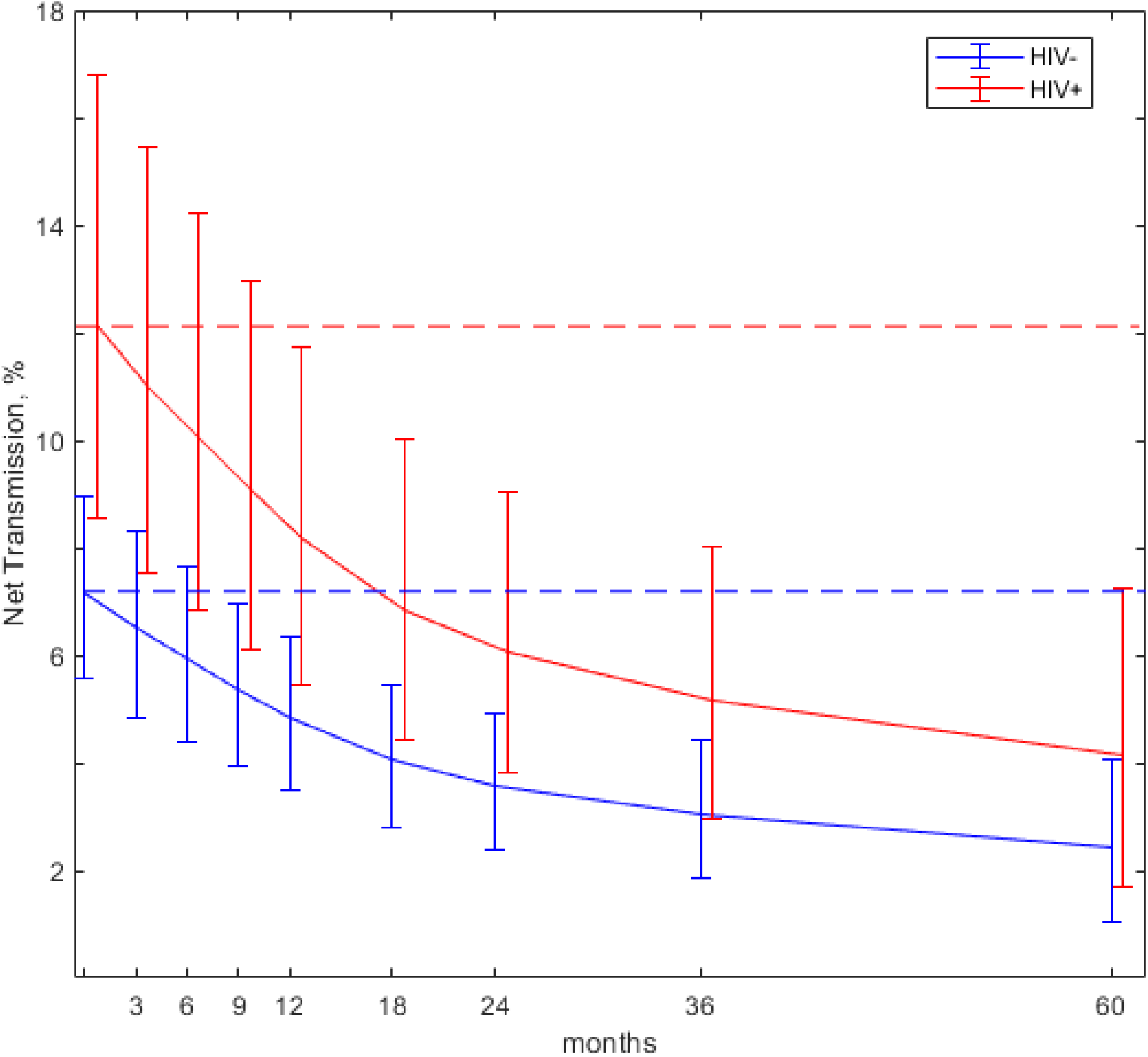
Overall vertical transmission (horizontal lines) and vertical transmission net of clearance at different ages: by mother’s HIV and weighted average of HIV- and HIV+.

Sensitivity analyses (Table 6) suggest that the main results, namely the overall VT rates and the proportion of infection by each route, are relatively insensitive both to model choice, and to assumptions about missing data. Goodness of fit statistics fail to distinguish between the alternative models (a difference of less than 3 is not regarded as meaningful), and none of the variations in modelling assumptions raise or lower key estimates by more than 5%, well within the statistical uncertainty of the preferred case model.

**Table 6.** Sensitivity analyses. Comparison of preferred model and alternatives. The preferred model is constrained interaction and assumes that 68.9% of the HIV -ve with unknown HCV RNA status are RNA positive, and 90.7% of the HIV +ve. Goodness of fit is posterior mean deviance.

|  | **Goodness of fit** | **Overall VT, % HIV-** | **Overall VT, % HIV+** | **Transmission by stage, %** |  |
| --- | --- | --- | --- | --- | --- |
| **Preferred model** |  |  |  |  |  |
| *Constrained interaction* | 741.6 | 7.2 | 12.1 | 24.8 | 9.3 |
| **Statistical uncertainty in preferred model** |  |  |  |  |  |
| *Lower (2.5%) credible limit* | - | 5.6 | 8.6 | 12.1 | 0.5 |
| *Upper (97.5%) credible limit* | - | 8.9 | 16.8 | 40.8 | 30.6 |
| **Model choice:** |  |  |  |  |  |
| *Main effect model* | 742.4 | 7.2 | 11.8 | 24.6 | 9.3 |
| *Simple interaction model* | 743.0 | 7.2 | 11.8 | 24.6 | 9.2 |
| **Proportion Low HCV viral load and proportion RNA -ve** |  |  |  |  |  |
| *Both odds lower by a factor of 1.6* | 742.2 | 7.4 | 12.2 | 24.6 | 9.1 |
| *Both odds higher by a factor of 1.6* | 740.9 | 7.0 | 12.1 | 24.8 | 9.4 |

## DISCUSSION

There has been little discussion of how the HCV VT rate should be calculated and reported: in particular, the question about how or whether clearance should be taken into account has received no attention. The few studies that investigated VT and clearance in the exact same data,[22, 23, 30] have reported VT rates of confirmed infection of 13.3%, 4.7% and 6.5% respectively, and 75%, 33.3% and 45% clearance before 5 years. These figures, however, are based on a total 32 infected children and do not account for unobserved clearance prior to age when infection status is ascertained.[24] The widely cited 25%-40% clearance rate[13] is based on data from the EPHN,[31] but the analysis included retrospectively recruited children, which is likely to have resulted in underestimates of clearance.[24]

The analysis here estimated 19.2 infections among the indeterminate and uninfected children, in addition to the 96 observed infections. The nominal transmission rate of 6.6% (Table 3) is in a population that includes 67.2% anti-HCV positive mothers with unknown PCR status, and is therefore lower than the VT rate that would be observed in a 100% HCV-RNA positive population. However, based on the VT rates in mono-infected and HIV-infected groups and the proportion HIV infected, it can be calculated that the 6.6% nominal rate is exactly what would be expected if 30.2% of the mothers with unknown HCV-RNA status were HCV-RNA negative and had a zero transmission rate. This accords with fact that 28% of the anti-HCV positive mothers were HCV-RNA negative in the EPHN cohort, and 31% in ALHICE.

When comparing results to previous literature, it is useful to consider VT rates in HIV uninfected and HIV co-infected mothers separately. The most recent meta-analysis of VT rates[2] reports 5.8% VT in HIV negative women. If we now apply 25%-40% clearance rates[31] (average 32.5%) to this, we predict that 3.9% of infants born to HCV-RNA positive mothers remain infected at 5 years. These figures can be compared to our estimated 7.2% overall transmission in mono-infected women and 2.4% net transmission at age 5 years. Thus, according to our analysis, the extent of VT is 24% higher than the accepted estimate, while the extent of chronic infection remaining at 5 years is 38% lower. Credible intervals should of course be taken into account (Figure 3).

**Figure 3.**
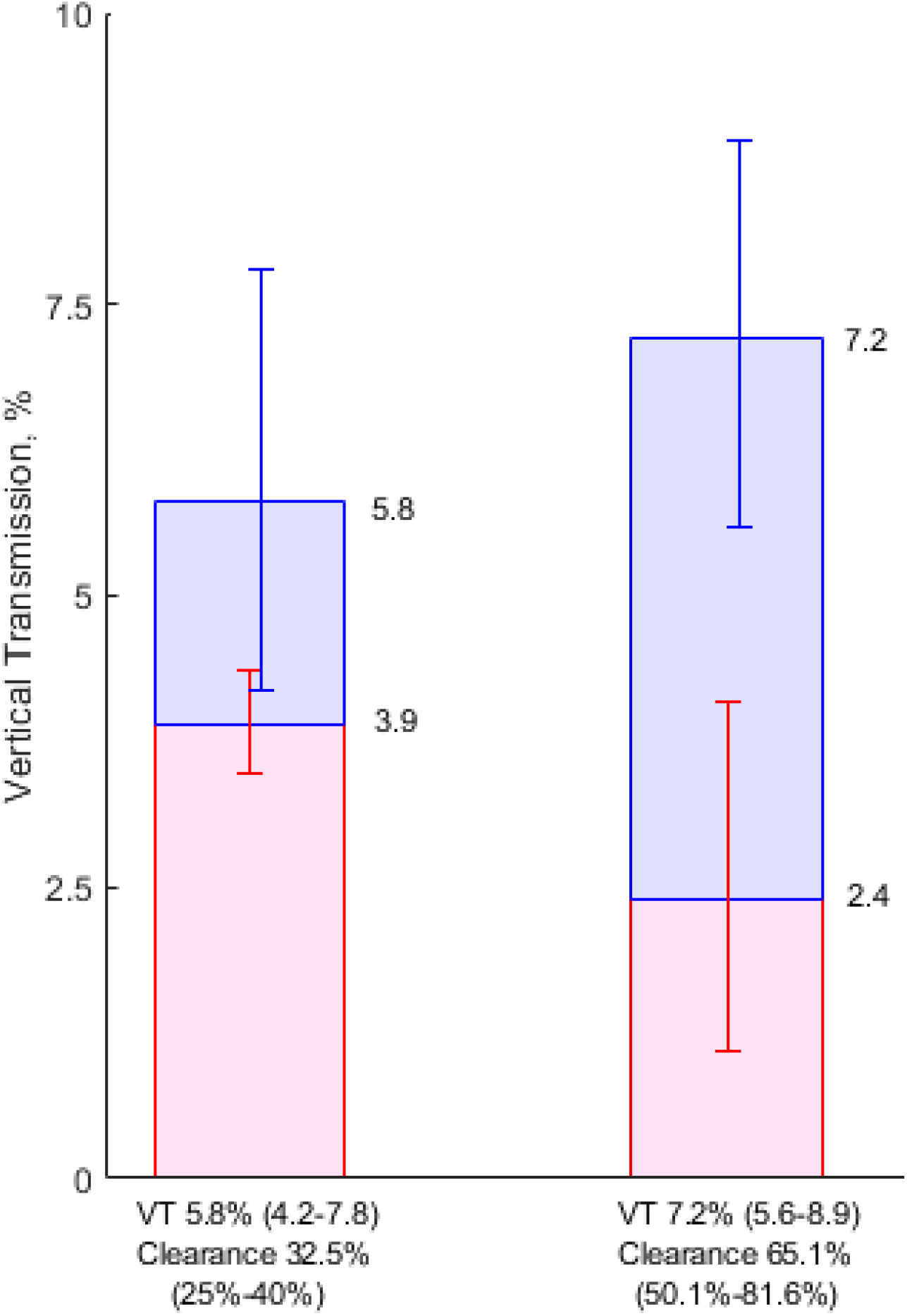
Left bar: VT rate of 5.8% (95%CrI: 4.2%-7.8%) in HCV mono-infected women[2], and spontaneous clearance 32.5% (25%-40%).[12, 16] Right bar: this study with VT rate of 7.2% (5.6-8.9) with 65.1% clearance (50.1%-81.6%). Blue segments: infection that clears within 5 years. Red segments: infection remaining after 5 years.

It appears that the meta-analysis VT rate of 5.8%, set against our estimate of 7.2%, represents a VT rate net of clearance at just under 6 months.[24] This accords with our finding that the average age at which uninfected children were first known to be uninfected was 5.1 months.

The analysis has a number of limitations. Much of the data was collected at a time when PCR tests were less accurate: various estimates of sensitivity and specificity of the tests used during this period have been made,[24, 32, 33] but, like most investigators, we have taken test results at face value for the sake of simplicity. This may have impacted on the classification of individuals as infected, uninfected and indeterminate, and on the assumed time to loss of anti-HCV in uninfected infants, time to clearance, and time to positive RNA in infected infants. Another drawback is the extent of missing data on mother’s viral load and RNA status. Although sensitivity analyses reveal that results are relatively robust against large changes in the assumed proportions RNA negative or with low viral load, the lack of data on this important risk factor has prevented us from investigating whether HIV and HCV-RNA status might impact transmission differently in utero or at delivery, or whether clearance rates themselves may be affected by risk factor. These questions do not appear to have been investigated previously.

Interpretation of any analysis based on confirmed infection is difficult because estimates of both VT and clearance rates will depend on the frequency and timing of testing. An alternative would be to base the clearance analysis not on “confirmed infection” but the presence of HCV-RNA.[24] This may make it easier to evaluate alternative protocols for early diagnosis of vertically-acquired infection, although the significance of viraemia in the absence of confirmed infection is not known.

Finally, our 12.2% estimate of the VT rate in HIV co-infected women may be of little contemporary relevance. Information on HIV treatment in two of our three cohorts was incomplete, but the majority of co-infected women would have been treated with the earlier generation of anti-retroviral drugs available up to 2003. Limited evidence from more recent European cohort studies including HIV/HCV co-infected women suggest substantially lower HCV VT rates, in the range 2.8%-5.9%,[34-36] in the context of high coverage with antiretroviral therapy and thus high levels of HIV suppression in pregnancy.

A major contribution of this paper is that it introduces methodology for simultaneously estimating overall VT rates and rates net of clearance. The novel element is the imputation of previously cleared infections among uninfected children, based on the age at which they were first known to be uninfected. This allows the investigator, for the first time, to accurately project the VT rate net of clearance at any age. Somewhat similar methodology, also applied in the present paper, has been used previously to impute the number of infections among indeterminates in studies of VT rates of both HCV[10] and HIV before PCR testing became widely available,[37-39] although in these studies the antibody loss curve was estimated simultaneously with the VT rate.

The second contribution is the findings on VT net of clearance. While they must be regarded as no more than provisional, given the shortcomings in the data, it is hoped that they will help inform trials of treatments in pregnancy to prevent vertical transmission. A recent phase I trial of 8 women suggested that sofosbuvir-ledipasvir in pregnancy was well-tolerated, achieved maternal cure and prevented VT[4] and further trials are under way.[5, 6] The recommended care of children exposed in utero is to delay diagnosis until 18 months and then refer anti-HCV positives for RNA confirmatory testing at 3 years prior to treatment.[13] It is questionable whether this strategy is viable where there is substantial loss to follow-up, as has been reported in infants born to HCV-infected women in the US,[18-20, 40] Our results may therefore also be relevant to evaluate alternative diagnostic and treatment strategies if and when treatments are licensed for use in children under 3 years.

## Data Availability

The data analysed for this paper was secondary. Individual patient data were requested from the studies listed in the manuscript and must be requested from the original research teams.

## Notes

### Author contributions

AEA conceived and carried out the analyses with the assistance of FG. AEA wrote the first and subsequent drafts of the paper. KS carried out the literature search and review. AEA, AJ, JC, and DMG were co-investigators on the HCVAVERT project, and AJ was the principal investigator. EC was a researcher on the HCVAVERT project. LP, EM-B, DMG and KB were senior or principal investigators on the 3 contributing studies: EPHN (European Pediatric HCV Network); ALHICE (Alpes-Maritimes, Languedoc, Haute Garonne Infection C chez l’Enfant); BPSU (British Pediatric Surveillance Unit). GI provided clinical input on hepatology and management of pediatric HCV. Curation of the original data files available to the project was the responsibility of LP and CT (EPHN), DMG, JC and KB (BPSU), and EM-B (ALHICE). Subsequent data processing was by FG and AEA. All authors critically reviewed and revised drafts as necessary and approved the final version for submission.

### Disclaimer

The funding sources did not have any influence on study design, data collection, analysis and interpretation of the data, writing of the report, or the decision to submit for publication.

## Acknowledgements

The authors express their gratitude to all those who have contributed to the EPHN, BPSU and ALHICE studies.

