## Supplementary Materials for "Overall vertical transmission of HCV, transmission net of clearance, and timing of transmission"

^1^ Population Health Sciences, University of Bristol Medical School, Whatley Road Bristol BS8 2PS. ^2^ MRC Clinical Trials Unit, University College London, 90 High Holborn, London WC1V 6LJ. ^3^ Population, Policy and Practice Research and Teaching Department, UCL Great Ormond Street Institute of Child Health, 30 Guilford Street, London WC1N 1EH. *^4^* London School of Hygiene and Tropical Medicine, Keppel Street, London WC1E 7HT. ^5^ Université Côte d'Azur, Public Health Department. Centre Hospitalier Universitaire de Nice, Rte St Antoine de Ginestière BP 3079. 06202 Nice cedex 2. ^6^ Children’s Health Ireland at Crumlin and Temple Street, Dublin, Ireland. ^7^ Meyer Children’s Hospital and Department Neurofarba, University of Florence, Viale Gaetano Pieraccini, 24, 50139 Firenze FI, Italy

*Joint last authors

**1. Definition of infection status**

Infants were considered *Infected* if they were anti-HCV positive after 18m or had at least two positive RNA tests; otherwise, they were *Uninfected* if they had any negative RNA results > 6 weeks, or if their last anti-HCV result was negative; otherwise, they were considered *Indeterminate.* Note that “Infection” should be interpreted as “ever-Infected” as an Infected infant may clear spontaneously and be uninfected by the end of follow-up. Similarly, those classified as “Uninfected” may have been infected and cleared before their infection was observed or confirmed.

Only one infant was deemed infected on the basis of anti-HCV beyond 18 months alone.

We assume that in all infected infants, RNA will be detectable by 6 weeks of age, based on previous literature.[1, 2] There is little to support the choice of 6 weeks as opposed to two or three months for this purpose: cases of late-appearing RNA have been reported in several cohorts and Thomas 1997[3] estimated that RNA was not detectable until after 3 months in 11% of infected children.

In our cohorts 7 infants who were considered infected based on at least two RNA positive tests had one or more negative RNA tests after 6 weeks and before RNA was detected. (Four cases were RNA-negative at 3 months, and two at 6 months, *before* having two or more positive RNA tests). Given the assumption that all infected children will be RNA+ve by 6 weeks, these 7 cases are anomalous. One possibility is that these cases represent post-natal intrafamilial infections, although this is considered rare.[4] We have included them as perinatally infected on the basis that the initial negatives after 6 weeks may have been false negatives caused by fluctuating levels of virus. A record review of these cases has appeared previously.[5]

**2. Statistical Methods**

The analysis is a Bayesian multi-parameter evidence synthesis,[6] a method which combines data informing different functions of a common underlying set of parameters.

***Model for timing of infection***

Transmission may take place at one of three stages “early *in utero*” (EiU), “late *in utero*” (LiU), and at delivery. For an infant in group
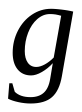
 (see below) the probabilities of transmission EiU, LiU, or at delivery, are
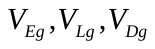
. These are to be interpreted as probabilities of transmission at each stage conditional on absence of transmission at a previous stage. Data is available on four functions of these parameters: probabilities of “ever” infection
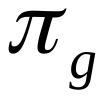
 in infants delivered by Elective Caesarean (ECS) and by Other Mode of Delivery (MoD), and probability of infection EiU conditional on vertical infection,
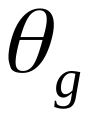
. The functions are:

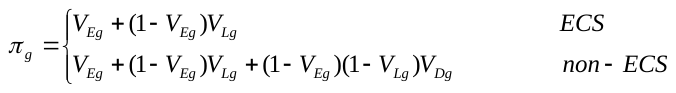

These functions of parameters are informed by the numbers of infected, uninfected and indeterminate children, and in the case of uninfected and indeterminate children their age at certain test results (see Likelihood contributions and probability of infection)

The probabilities of EiU transmission conditional on infection, following ECS and following non-ECS deliveries are:

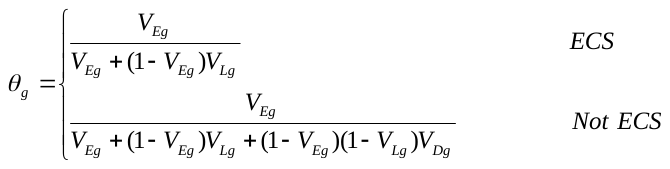

These functions of parameters are informed by the binomial data on the proportion of infants tested in the first 3 days who were HCV-RNA positive.

***Risk factors, risk combination models, and prior distributions***

Taking
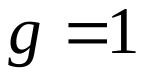
 as representing the EPHN dataset, HIV-ve, Low Viral Load group, the risk of transmission in other groups is

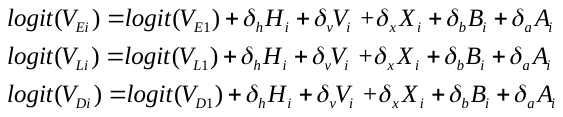

Where
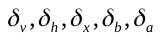
 are coefficients (log odds ratios), and
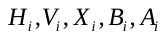
 are indicators for HIV +ve, High Viral Load, the interaction (HIV+ve *and* High Viral load), the BPSU study, and the ALHICE study.

The stage-specific transmission rates in the EPHN study in the lowest risk group,
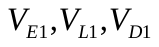
were given weakly informative priors, to stabilize computation and rule out implausibly high or low posteriors:

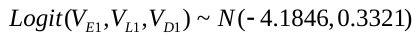

This implies an *a priori* stage-specific probability of transmission with mean 0.045, median 0.015, and 95% interval 0.0005 to 0.24.

Three models were fitted. In the *Simple Interaction* model uninformative priors are placed on all coefficients:
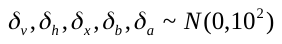
. The *Main Effects* model has the same priors except that
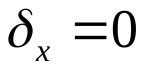
. In the *Constrained Interaction* model
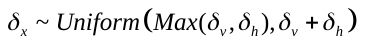
. This has the effect of ensuring that the combined effect of high viral load and HIV+ cannot be less that either factor alone, and cannot be greater than their sum (on the log scale). The simple interaction. and Main Effect models were examined as sensitivity analyses.

***Missing data***

A high proportion of data on mother’s HCV RNA status and mother HCV viral load was missing. We assumed that the proportion of RNA-unknown mothers who were RNA-positive was the same as the proportion of RNA-known mothers who were RNA-positive, and that the proportion of viral load-unknown who had low viral load was the same as the proportion of viral-load-known women with low viral load. These proportions were extracted from the EPHN data, and applied to both the EPHN and BPSU datasets. Given those assumptions, the expected probability of transmission in the missing data groups can be expressed as simple functions of the probabilities in groups with known risk factors, as follows:

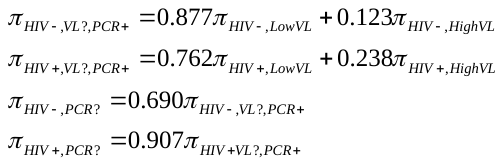

The assumed proportions were subjected to sensitivity analysis in which the odds were both increased or decreased by an odds ratio of 1.6.

***Likelihood contributions and probability of infection***

The data structure comprises four independent datasets, each of which informs different functions of the underlying of “basic”[6, 7] parameters. The basic parameters are:
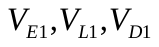
 and
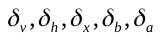
.

The first dataset informs the probability of detectable RNA in infected infants tested in the first 3 days, conditional on risk group. This is a grouped binomial likelihood:
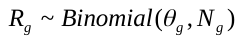
. The remaining three datasets comprise the Infected, Indeterminate, and Uninfected infants respectively, and these can be considered together.

If
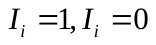
 indicate that individual is infected and not infected respectively, then the likelihood contribution of each
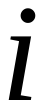
with observed data
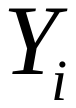
 is :
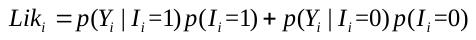

The likelihoods for each *type* of observation are set out in Table S1, along with the probability of infection conditional on the outcome pattern.

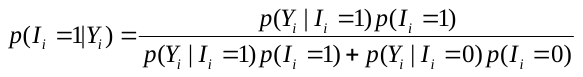

The different observation types are:

1. *Infected*
2. *Uninfected,* subdivided into two groups:

(a) The probability they were infected and then cleared increases with the age of the first RNA or anti-HCV test that shows they were uninfected

(b) Some also have RNA -ve tests in the first 6 weeks. This lowers the probability they were infected – the more so as the age (<6w) increases

3. *Indeterminate*, sub-divided into three groups:

(a) Those with an anti-HCV positive test < 18m. The probability they are infected increases with age at the last anti-HCV positive < 18 months

(b) Those with an RNA negative test < 6w. The later the test the lower the probability of infection.

(c) Those with *both* an anti-HCV test *and* an RNA negative test under 6 weeks.

The likelihoods for each group (Table S1) depend on the following functions, which have been estimated in advance using data from the same three cohorts. All available data was used, including data on individuals with missing data on risk factors.

-
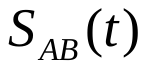
, the “antibody loss curve”: this is the probability of still being anti-HCV +ve at
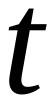
 in an uninfected infant.
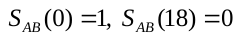
.
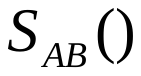
was estimated from interval censored and right censored data on 1697 uninfected infants. A normal time-to-event curve was fitted producing an estimated mean 8.44m (95%CrI 8.21 - 8. 67) and standard deviation 3.82m (95%CrI 3.65 - 4.00).
-
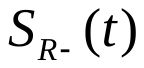
 is the probability that an infected infant who is not already RNA+ at birth remains RNA -ve at
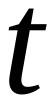
. Time to RNA +ve is assumed to be normally distributed.
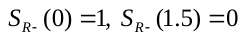
.
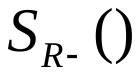
was estimated from records on 37 confirmed infected infants whose first RNA test was negative. A normal curve was fitted to interval censored data on age at first RNA +ve and age at the immediately preceding RNA -ve. The curve was constrained so that the estimated cumulative proportion with detectable RNA would be 99.9% at age 45 days. The resulting curve had mean 0.83m (0.69 - 0.92) and standard deviation 0.21m (0.17 – 0.27)
-
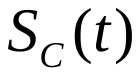
, the clearance of infection curve. This is the probability that an infected infant remains infected (RNA +ve) at time
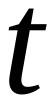
. This function was estimated previously,[5] based on a natural cubic spline model fitted to the interval censored and left truncated data on 106 infants with confirmed infection. An estimated 65.9% (95%CrI: 50.1-81.6) of confirmed infections cleared by age 5 years.

The three curves were discretized to decrease computation time, and they are shown in that form in Figure 1 Panels A1, B1, C1 in the main text.

***Bayesian Estimation***

WinBUGS 1.4.3 was used for estimation. Convergence occurred within 20,000 iterations, based plots of the Gelman-Rubin statistic. Posterior summaries were based on 30,000 samples from each of three chains, after a burn-in of 30,000 iterations.

**Table S1.** Likelihood contributions

| **Outcome** | **Probability of outcome if infected** | **Probability of outcome**  **if not infected** | **Contribution to likelihood** | **Probability infected, given outcome** |
| --- | --- | --- | --- | --- |
| *Infected* | 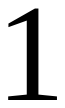 | 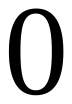 | 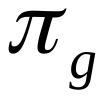 | 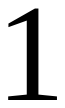 |
| *Uninfected* (a): first RNA -ve test at 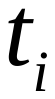>6w | 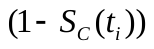 | 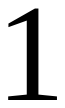 | 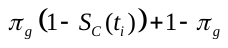 | 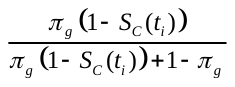 |
| *Uninfected* (b): with last RNA-ve <6w at 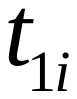AND a first negative test >6w at 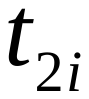 | 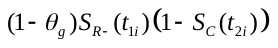 | 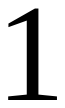 | 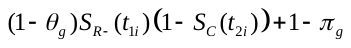 | 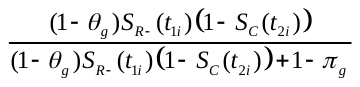 |
| *Indeterminate* (a): last anti-HCV +ve, 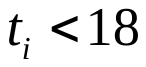 | 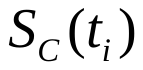 | 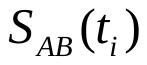 | 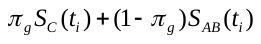 | 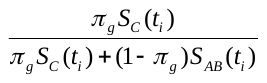 |
| *Indeterminate* (b): last RNA-ve 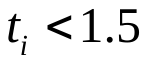 | 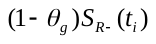 | 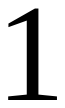 | 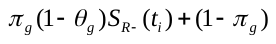 | 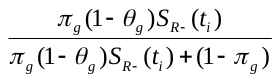 |
| *Indeterminate* (c): last RNA-ve, 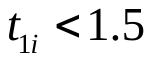 AND last anti-HCV +ve, 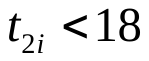 | 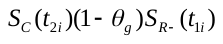 | 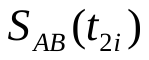 | 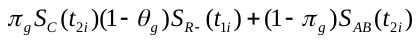 | 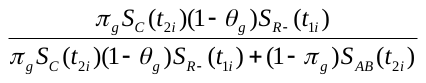 |
